# SARS-CoV-2 transmission during team-sport: Do players develop COVID-19 after participating in rugby league matches with SARS-CoV-2 positive players?

**DOI:** 10.1101/2020.11.03.20225284

**Authors:** Ben Jones, Gemma Phillips, Simon PT Kemp, Brendan Payne, Brian Hart, Matt Cross, Keith A Stokes

## Abstract

**Objectives:** Evaluate the interactions between SARS-CoV-2 positive players and other players during rugby league matches, to determine the risk of in-game SARS-CoV-2 transmission.

**Design:** Observational.

**Setting:** Super League rugby league during four matches in which SARS-CoV-2 positive players were retrospectively found to have participated (2^nd^ August and 4^th^ October 2020).

**Participants:** 136 male elite rugby league players: eight SARS-CoV-2 positive participants, 28 identified close contacts and 100 other players who participated in any of the four matches.

**Main Outcome measures:** Close contacts were defined by analysis of video footage for player interactions and microtechnology (GPS) data for proximity analysis. Close contacts and other players involved in the matches becoming positive for SARS-CoV-2 by RT-PCR within 14 days of the match were reported.

**Results:** The eight SARS-CoV-2 positive players were involved in up to 14 tackles with other individual players. SARS-CoV-2 positive players were within a 2 m proximity of other players for up to 316 secs, from 60 interactions. One identified contact returned a positive SARS-CoV-2 result within 14 days of the match (subsequently linked to an outbreak within their club environment, rather than in-match transmission), whereas the other 27 identified contacts returned negative SARS-CoV-2 follow up tests and no one developed COVID-19 symptoms. Ninety-five players returned negative and five players returned positive SARS-CoV-2 RT-PCR routine tests within 14 days of the match. Sources of transmission in the five cases were linked to internal club COVID-19 outbreaks and wider-community transmission.

**Conclusion:** Despite a high number of tackle involvements and close proximity interactions between SARS-CoV-2 positive players and players on the same and opposition teams during a rugby league match, these data suggest that in-game SARS-CoV-2 transmission is limited during these types of team sport activities played outdoors.

## INTRODUCTION

The COVID-19 pandemic caused professional and community level team-sports around the world to be postponed.[1] COVID-19 is caused by Severe Acute Respiratory Syndrome Coronavirus 2 (SARS-CoV-2)[2] and transmitted from human-to-human by multiple means, namely, by respiratory droplets, aerosols, and fomites.[3,4] As the community COVID-19 prevalence rises high, the likelihood of a COVID-19 positive participant entering a sporting environment also rises. Even without considering the risk of crowds,[5] given the close proximity of participants and increased respiration rate due to the demands of exercise,[6] team sports pose a potential risk for human-to-human transmission.

The implementation of a ‘Test and Trace’ system, reduces the risk of wider community spread.[7] If a participant of a team sport test positive for SARS-CoV-2 following an activity, those deemed to be a close contact should isolate for 14-days.[8,9] The mental, physical, economic and societal impact of a 14-day isolation period should not be underestimated,[10–12] therefore contact tracing should be done with appropriate precision in order to mitigate against the potential adverse effects of both under and over isolation. The return of community sport requires decisions to be made regarding who needs to isolate, should a participant test positive for SARS-CoV-2. These decisions should ensure community transmission in minimised, whilst considering the negative impact of unnecessary isolation. Managing and mitigating the risk of virus transmission should be balanced with the benefits of sport on physical and mental health.[13]

Rugby league includes frequent contacts (e.g., tackles)[14] and close proximity interactions between players, similar to other rugby and football codes.[15] The repeated close contacts between multiple participants during a match represent theoretical opportunities for transmission of SARS-CoV-2, given droplets, aerosols and fomites are the primary means of transmission.[3]

Professional rugby league (i.e., Super League competition) restarted on the 2^nd^ August 2020, with a number of risk management and mitigation factors implemented.[16] These included law modifications (e.g., removal of the scrum), routine weekly reverse transcriptase polymerase chain reaction (RT-PCR) screening for the presence of SARS-CoV2 RNA, created in response to a SARS-CoV-2 infection,[17] daily self-reporting of potential COVID-19 symptoms, in addition to other policies relating to the biosecurity of training (e.g., cleaning procedures) and match venues (e.g., no spectators), similar to other sports.[18]

Since the start of the Super League a number of players have tested positive for SARS-CoV-2 at the next routine RT-PCR screening following a match. Consequently, *‘Contacts’* have been identified using predetermined criteria.[16] Rugby league provides an interesting case study to understand the risk of SARS-CoV-2 transmission during team sports, which may help to inform test and trace activities in other sports. This study aims to i) describe and evaluate the interactions between players who tested positive for SARS-CoV-2, and other players during matches, and ii) document the time course of testing and monitoring of identified contacts, who were required to isolate.

## METHODS

### Study Overview and Ethics

Rugby league matches returned on the 2^nd^ August 2020 following the COVID-19 enforced shutdown. Thirty-six matches were played during this observational period (1^st^ July [start of RT-PCR screening] - 4^th^ October 2020). Each match directly involves 34 players (17 players on each team; 13 starting and 4 as interchange) and one on-field match official and two touch judges. Of the 36 matches, there were four occasions when eight players (CoV1-CoV8) from four Super League teams, tested positive for SARS-CoV-2 either during routine RT-PCR screening or following the development of symptoms, and contacts were identified (C1-C28). All participants were male professional rugby league players. All participants were within a ≤7-day RT-PCR screening cycle, thus returned a negative test in the 7 days prior. Teams had autonomy on which day they undertook RT-PCR screening, which were typically at the start of their training week. SARS-CoV-2 positive players were deemed at risk of shedding infectious virus during a match, where the symptom onset or the test occurred within 48 hrs of a match. Consequently, contact tracing was carried out on four Super League matches, resulting in 28 players isolating for 14 days from exposure, in accordance with Public Health England guidance. The number of positive cases and identified contacts from each match was; 5 and 12, 1 and 3, 1 and 9, and 1 and 4, respectively. Ethics approval (Reference; 73648) was obtained from Leeds Beckett University local ethics committee.

In total, for rugby league players within this RT-PCR monitoring pool, 29 players tested positive for SARS-CoV-2 from 4572 tests, between 1^st^ July and 4^th^ October 2020. Two ‘clusters’ were observed (Club 1 [match 1]; *n* = 9 players, 3 staff, Club 2 [match 2]; *n* = 4 players, 2 staff), indicating evidence of transmission within a sporting environment, in addition to wider community transmission into rugby league.

Only the eight players deemed to be shedding infectious virus (i.e., a positive SARS-CoV-2 RT-PCR screening test within 48 hrs from a match or COVID-19 symptoms within 48 hrs from a match followed by a subsequent positive SARS-CoV-2 RT-PCR test), which also resulted in other players being identified as a contact from a match were included in this study. Other SARS-CoV-2 positive players did not participate in a match within a timeframe when they would have been shedding infectious virus (e.g., returned a positive SARS-CoV-2 RT-PCR screening test or developed COVID-19 symptoms greater than 48 hr following a match; *n* = 20) or participated in a match but did not result in a contact being identified (*n* = 1).

### Identification of COVID-19 Positive Players

Players undertook weekly SARS-CoV-2 RT-PCR testing and undertook daily COVID-19 symptom monitoring. Swabs were taken from the nasopharynx and oropharynx by trained health care professionals. All testing was via an ‘end-to-end’ offered by the testing provider and undertaken within nationally accredited laboratories. Target genes were N, S, ORF1ab. Most positive (CoV1-CoV5, CoV7-CoV8) and negative tests were analysed in the same laboratory (SARS-CoV-2 CE-IVD rt-PCR Test, Oncologica UK Ltd, Cambridge, England). If a player returned a positive test, their respective cycle threshold (Ct) values (where available) for each gene (N, S, ORF1ab) and clinical presentations were reviewed by an independent consultant virologist to ensure this was a *‘true positive’*. Players’ symptoms were monitored by their team physician. The monitoring covered all typical COVID-19 symptoms (e.g., cough, fever, smell and taste disturbances, difficulties breathing).[19]

### Identification of Contacts

If a positive SARS-CoV-2 screening test was returned within 48 hrs of a match (Figure 1; players CoV1-CoV5, CoV7), or development of symptoms consistent with COVID-19 were within 48 hrs of a match, with a subsequent positive SARS-CoV-2 test within 48hrs (Figure 1; players CoV6, CoV8), it was assumed the player was shedding infectious virus during the match [20], and contacts were then identified. This included players on the same and opposition team.

**Figure 1.**
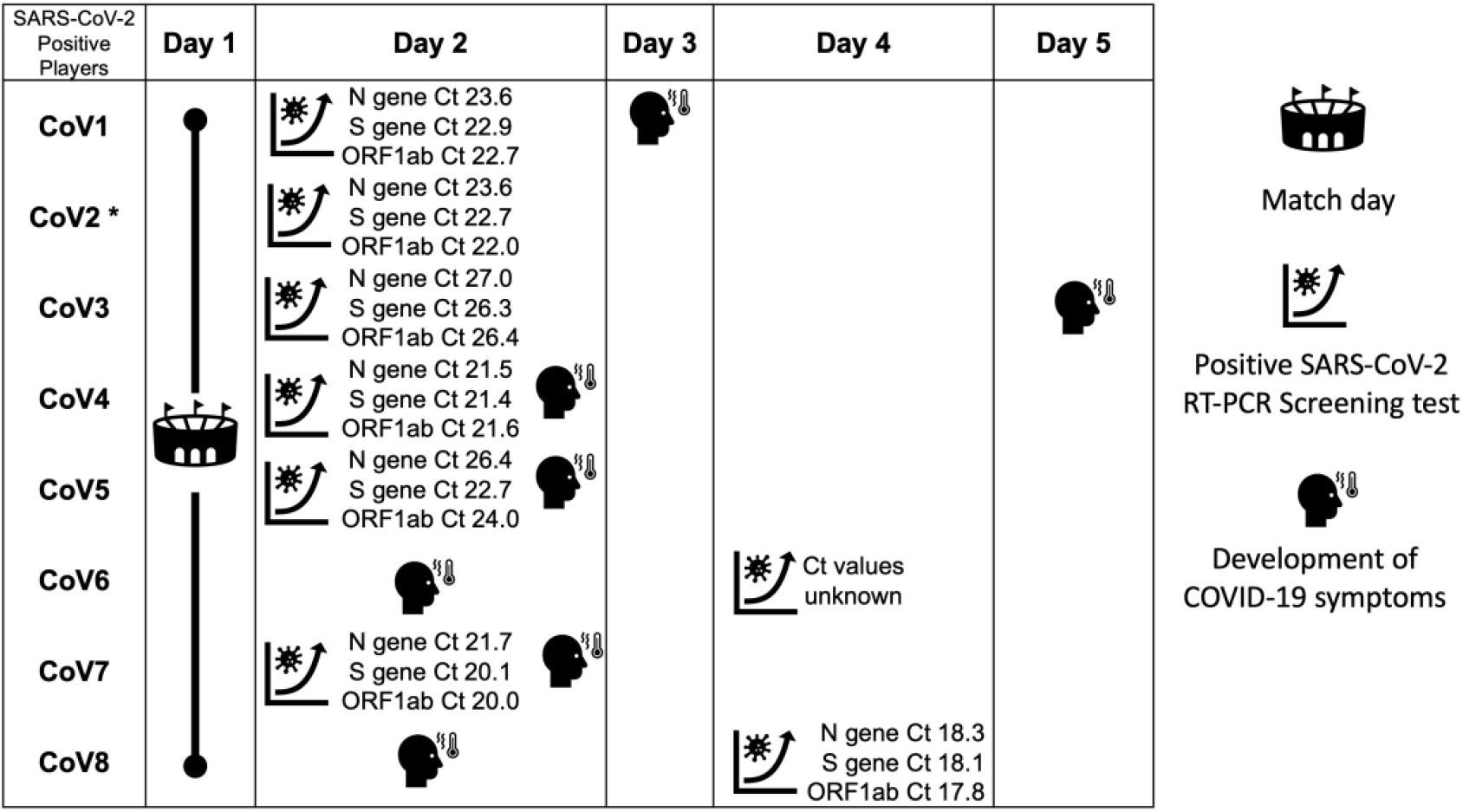
Time course of positive test, test characteristics and development of symptoms for SARS-CoV-2 positive players. *CoV2 did not develop symptoms. Positive SARS-CoV2 RT-PCR screening depicted on day of test, not day of result.

Contacts were defined based on definitions agreed by experts in public health and sports medicine.[16] Between kick-off and full-time, any player from the match who was within 1 m, face-to-face for ≥3 secs was deemed a contact and consequently required to isolate for 14 days. This definition was used as other broader public health definitions (e.g., <2 m for >15 min,[21]) were not deemed to capture the close contact face-to-face and fleeting encounters that occur during team sports.

Match statistics for SARS-CoV-2 positive players (identifying tackle involvements, as either a ball carrier [i.e., attacker] or tackler [i.e., defender]) were provided by a commercial match statistics provider (Opta, Leeds, United Kingdom) who analysed the match video footage. The match was then analysed by the same experienced Performance Analyst to identify video footage of tackles involving SARS-CoV-2 positive players. Tackles and ball-carries were reviewed numerous times in slow motion to determine which tackles involved a <1m, face-to-face interaction for ≥3 secs with another player. Clips were then reviewed by a second reviewer to confirm or reject the identified contacts. Identified contacts were then informed by the national governing body and their respective club and required to isolate for 14 days.

### Tackle Involvements and Player Proximity

In addition to the identification of contacts (i.e., <1m, face-to-face for ≥3 secs; C1-C28), to determine tackle involvements, a matrix was produced using the match statistics to determine how many times SARS-CoV-2 positive players were involved in tackles with other players (opposition and same team, as some tackles involve more than one defender).

The Super League operates a league-wide microtechnology project (Project SL-Catapult), whereby all teams are supplied with the same devices (Optimeye S5, Catapult Sports, Melbourne, Australia).[22] When both teams wore microtechnology devices (matches 1, 3 and 4), raw longitude and latitude data from global positioning system (GPS) were analysed to determine the number of occasions and the duration of time SARS-CoV-2 positive players were ≤2 (±1) m of other players [23]. The number of occasions and the duration of time within ≤2 (±1) m, were analysed for different dwell times (e.g., duration of close proximity encounter), to establish the nature of the interactions. One team did not wear their microtechnology units, thus player proximity was not determined for match 2.

### Patient and Public Involvement

All data included in this study were collected routinely by teams or the National Governing Body (Rugby Football League) as part of the Super League competition. There was no patient or public involvement in the study.

## RESULTS

### SARS-CoV-2 Positive Player Test Characteristics

The positive RT-PCR test characteristics (where available) and the timing of development of symptoms in relation to the match and are shown in Figure 1. Players developed COVID-19 symptoms prior to (*n* = 2; CoV6, CoV8), on the day of (*n* = 3; CoV4, CoV5, CoV7) or following (*n* = 2; CoV1, CoV3) their positive SARS-CoV-2 RT-PCR test. One player did not develop COVID-19 symptoms (CoV2).

### Interaction between SARS-CoV-2 Positive Players and other players during a Match

The SARS-CoV-2 positive players (CoV1-CoV8) and their interactions with other players during the match are shown in Figures 2-4.

**Figure 2.**
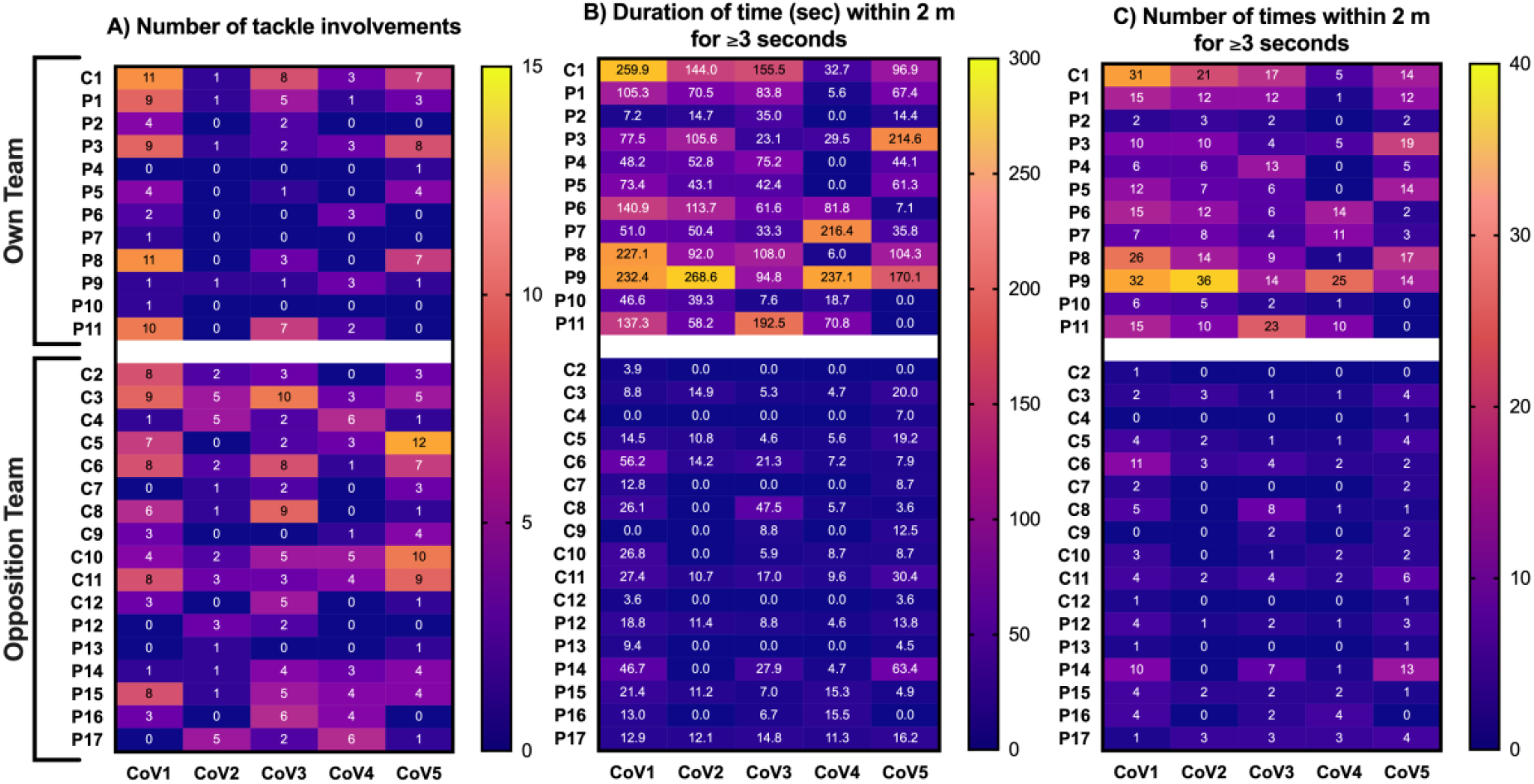
Number of tackle involvements and close proximity (<2 m) interactions between SARS-CoV-2 positive players (CoV1 – CoV5), identified contacts (C1-C12), and other players (P1-P17) during rugby league match 1.

**Figure 3.**
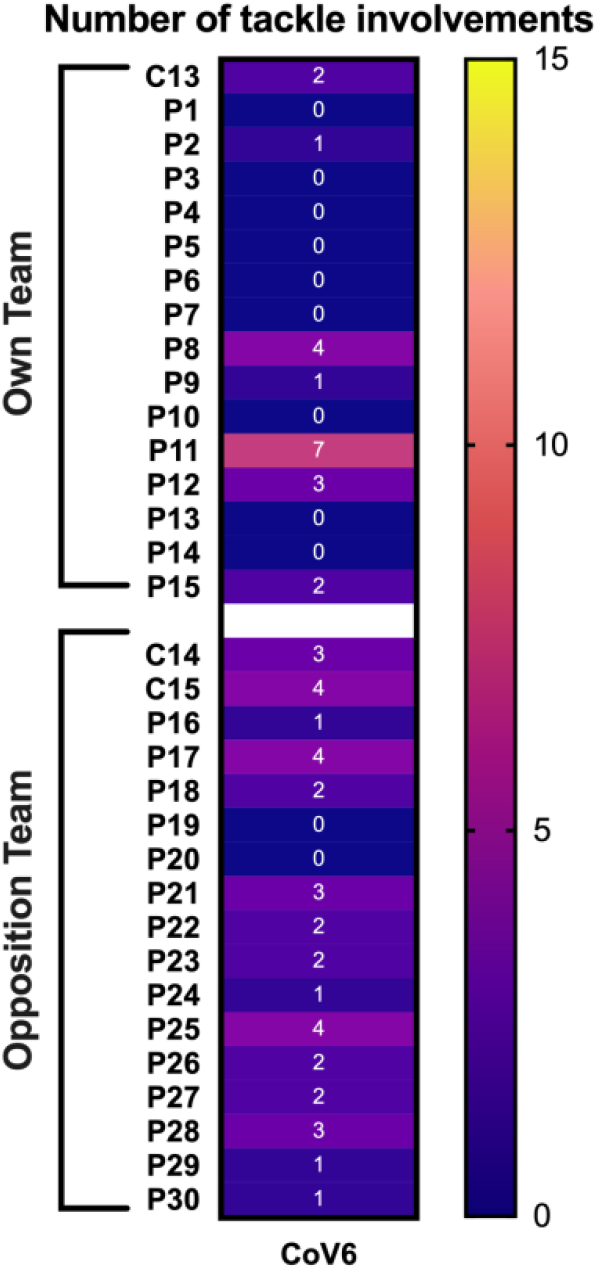
Number of tackle involvements between SARS-CoV-2 positive player (CoV6), identified contacts (C13-C15), and other players (P1-P30) during rugby league match 2.

**Figure 4.**
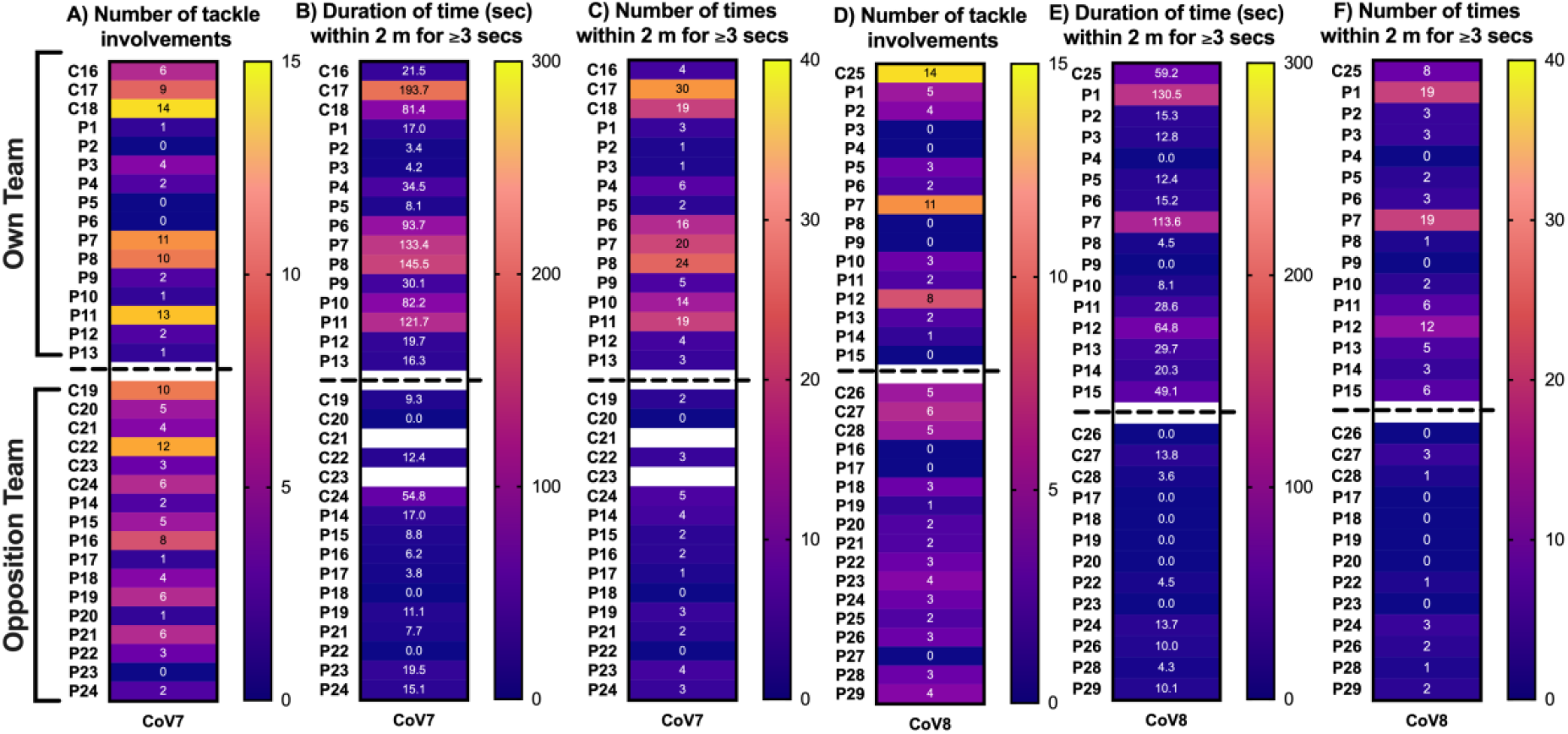
Number of tackle involvements and close proximity (<2 m) interactions between SARS-CoV-2 positive players, identified contacts, and other players during rugby league match 3 (Fig 4A-4C; CoV7; C16-C24, P1-P24) and match 4 (Fig 4D-4F; CoV8; C25-C28, P1-P29). C21 and C23 had a GPS unit error.

Identified close contact tackles (i.e., <1 m, face-to-face for ≥3 secs) were observed between CoV1 and C1, C6, C9, C12; CoV2 and C2; CoV3 and C6, C8; CoV4 and C4, C11; CoV5 and C3-C5, C7, C10, C11; CoV6 and C13-C15; CoV7 and C16-C24; CoV8 and C25-C28.

The number of tackle involvements between SARS-CoV-2 positive players (i.e., CoV1-CoV8) and identified contacts (i.e., C1-C28) were ≥10 on eight occasions (Figures 2A, 3, 4A, 4D). SARS-CoV-2 positive players were also involved in ≥10 tackles with non-contacts on six occasions. Based on the GPS analysis, SARS-CoV-2 positive players were within 2 m for ≥3 secs for 70.2 ± 67.8 secs of players on their own team and 9.5±21.1 secs of players on the opposition team (Figures 2B, 4B, 4E). SARS-CoV-2 positive players were within 2 m for ≥3 secs on 9.5 ± 8.3 occasions for players on their own team and 1.9 ± 2.3 occasions for players on the opposition team (Figures 2C, 4C, 4F).

Close proximity interactions between SARS-CoV-2 players (CoV1-CoV5, CoV7-CoV8) and identified contacts (C1-12, C16-C20, C22, C24-C28) are shown in Figure 5A, for different epochs. Interactions were typically <5 secs. Sixty-three percent of all interactions were <3 secs. CoV1, CoV2, CoV3, CoV7 and CoV8 had an interaction with a contact for ≥20 secs (Figure 5B).

**Figure 5.**
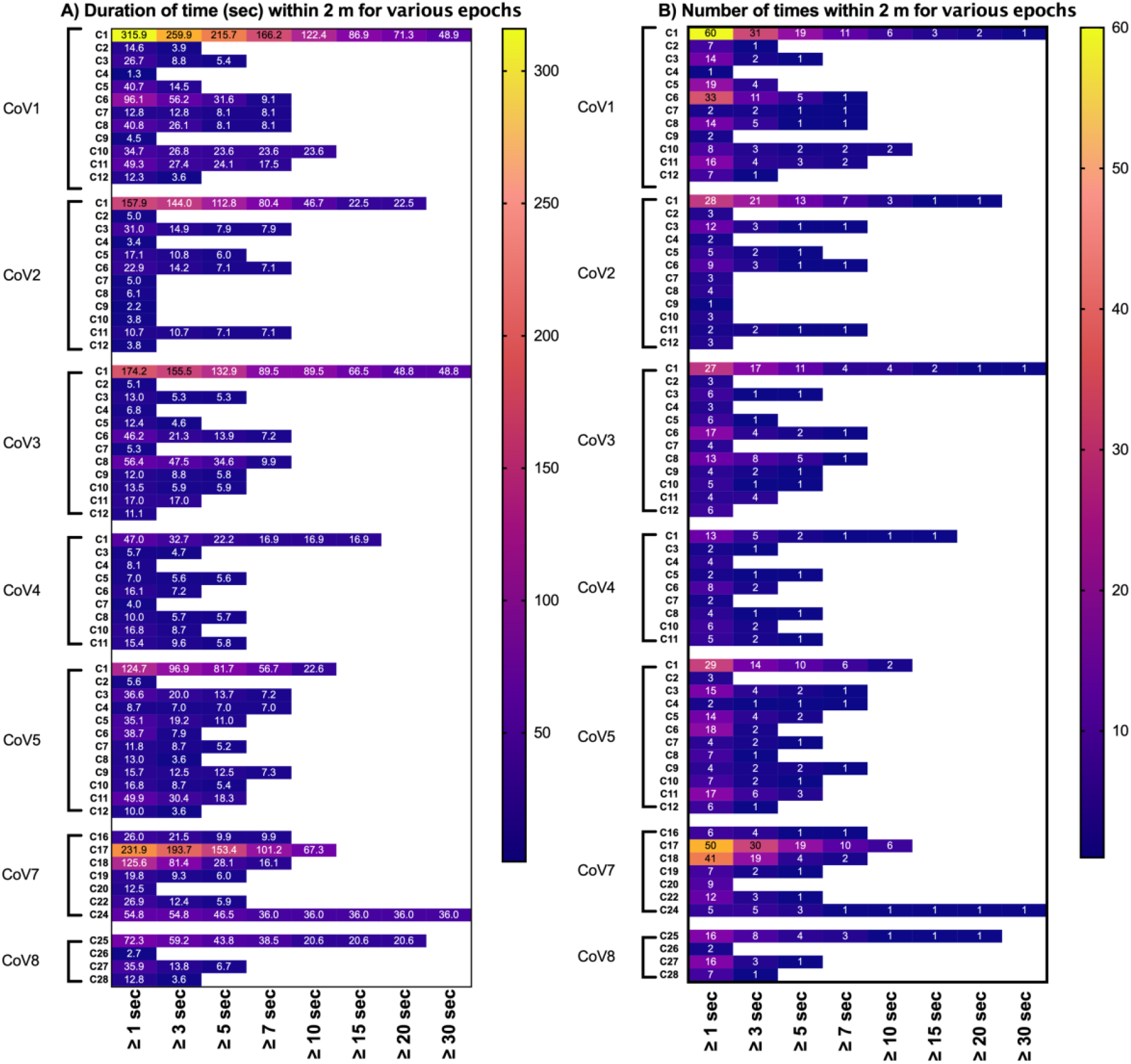
Duration (A) and number (B) of close proximity (<2m) interactions between SARS-CoV-2 positive players and identified contacts, for various epochs. C21 and C23 had a GPS unit error.

### Testing and Symptom Monitoring of Contacts

During the 14-day isolation period, contacts completed their normal daily self-reporting of potential COVID-19 symptoms screening and had 1–4 SARS-CoV-2 RT-PCR tests, to determine if they contracted the virus. Contacts also had SARS-CoV-2 RT-PCR tests on day 16 or 17 (Figure 6).

**Figure 6.**
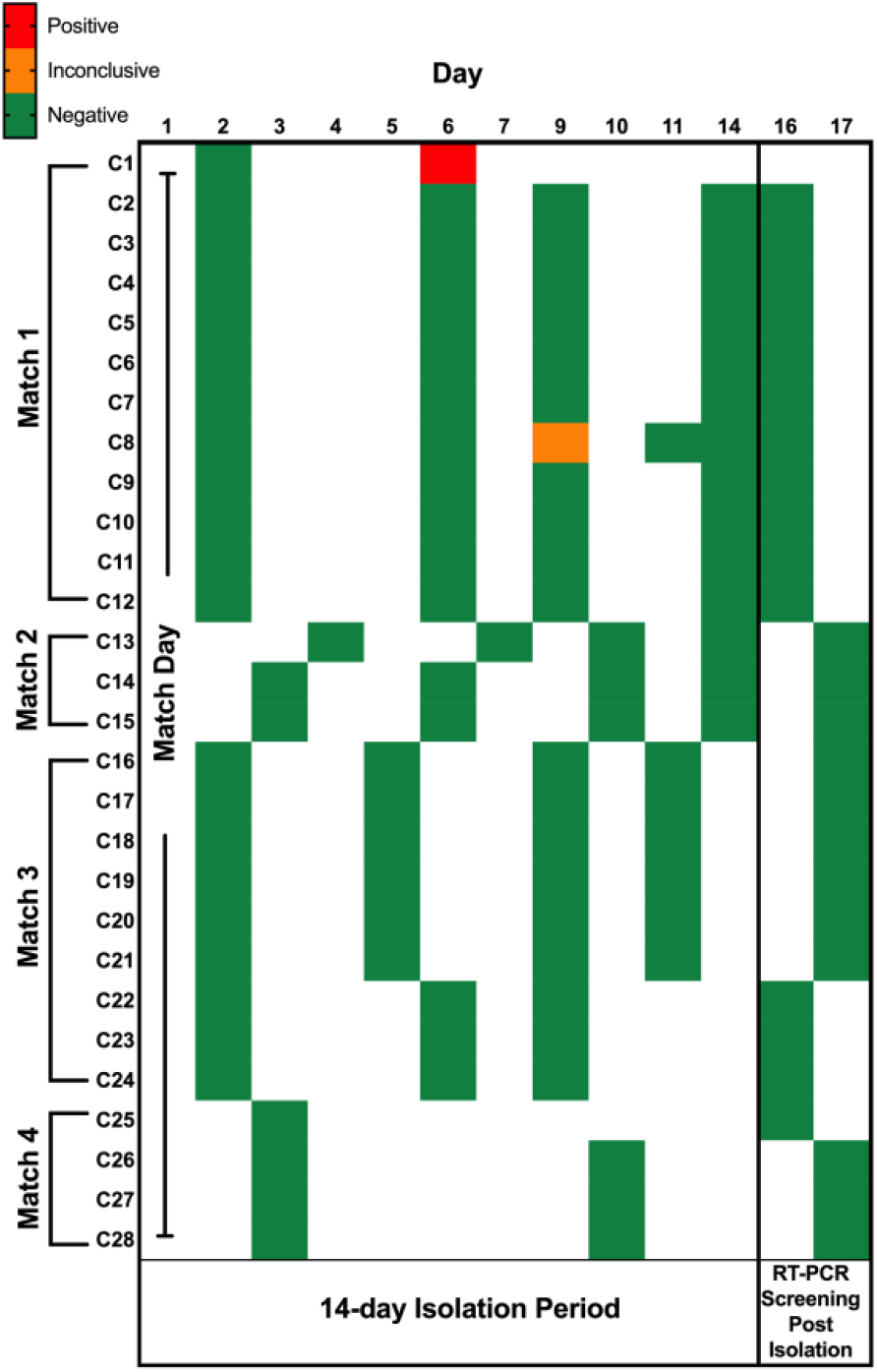
Time course of testing for contacts (C1-C28) required to isolate, due to interactions with SARS-CoV-2 positive players during a match.

During this period of observation, C1 tested positive for SARS-CoV-2, whereas all other contacts (C2-C28) returned negative SARS-CoV-2 RT-PCR tests. Out of the other 100 players participating in the matches with SARS-CoV-2 positive players (i.e., non-contacts or non-SARS-CoV-2 positive players; Figures 2, 3 and 4), five players returned positive SARS-CoV-2 RT-PCR screening results and 95 players returned negative SARS-CoV-2 RT-PCR screening results (Table 1).

**Table 1.**
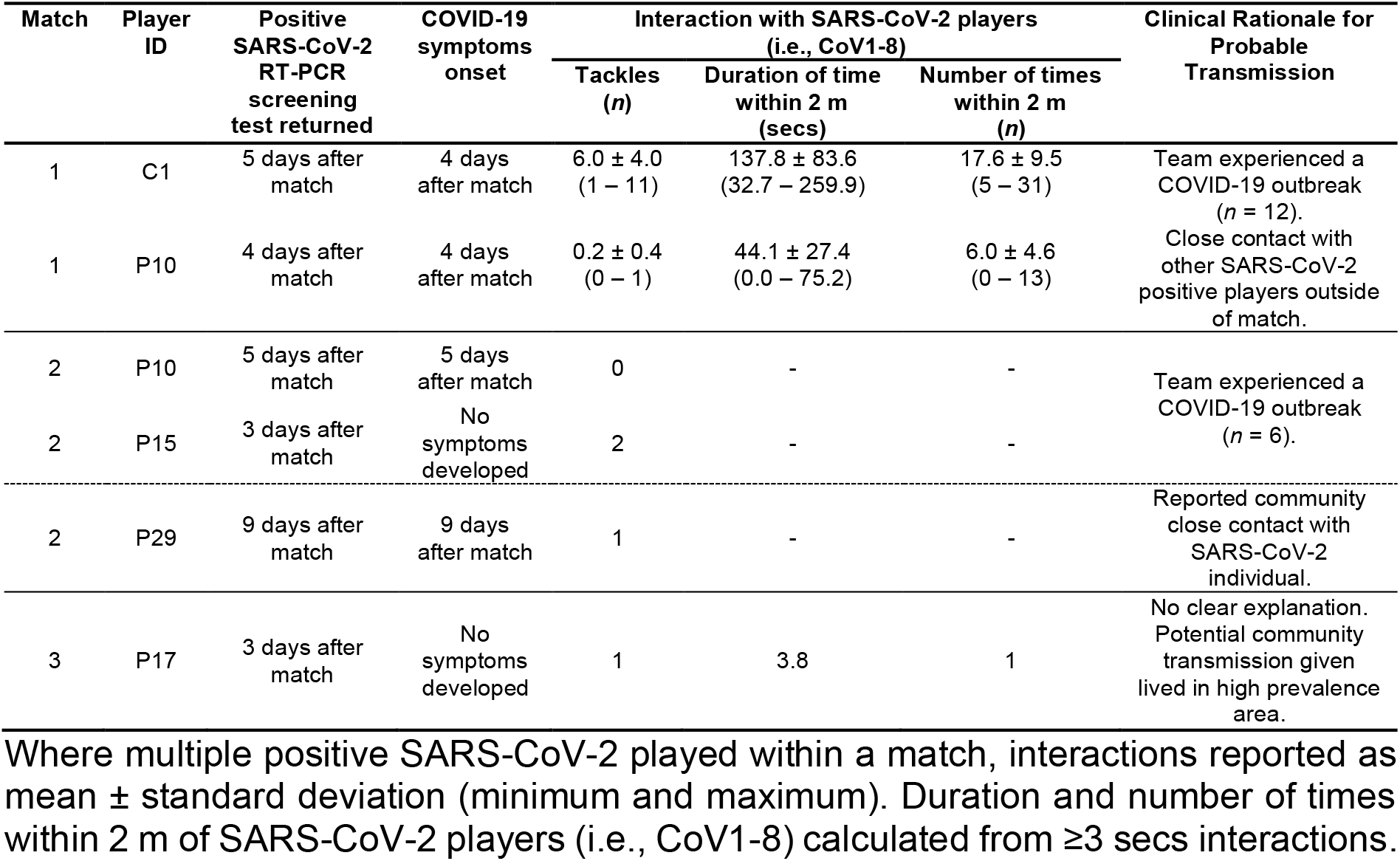
Match, Player ID, positive test and symptom timelines, interactions with SARS-CoV-2 positive players, and clinical rationale for probable transmission

On review, SARS-CoV-2 positive cases for C1 and P10 (match 1) were linked to a COVID ‘outbreak’ in the club and a further two players (who did not play in the match) and three staff tested positive for SARS-CoV-2 at this club (match 1). Players (C1 and P10) also reported non-training close contacts with SARS-CoV-2 players within the club environment (Table 1). C1 had the greatest duration and number of close proximity interactions with SARS-CoV-2 positive players during the match in comparison to other identified contacts, whereas P10 only involved in 1 tackle with CoV1 during the match (Figure 2A).

P10 and P15 (match 2) were also considered to be part of a COVID-19 outbreak within their club environment (Table 1). P29 (match 2) was considered to be wider-community transmission, due to the reporting of a community close contact. P29 also returned a negative SARS-CoV-2 RT-PCR screening test between match 2 and returning a positive SARS-CoV-2 RT-PCR screening. P17 (match 3) provided no clear explanation for transmission, although lived in an area of high COVID-19 prevalence, thus was considered to be wider-community transmission.

All match officials (*n* = 40) involved in Super League during this observational period returned negative SARS-CoV-2 RT-PCR screening tests (from 312 weekly screening tests). This includes following the four matches when positive SARS-CoV-2 players were participating.

## DISCUSSION

Professional and community team sports are returning during the COVID-19 pandemic, although transmission during these activities is relatively unknown. For the first time, this study presents detailed player-to-player interactions of eight players who participated in rugby league matches whilst infectious with SARS-CoV-2. Twenty-eight players were identified as contacts due to their interactions with SARS-CoV-2 positive players and were required to isolate. SARS-CoV-2 positive players were involved in up to 14 tackles and spent up to (approx.) 5 mins within 2 m of identified contacts during the match. Twenty-seven identified contacts returned negative, and one identified contact returned a positive RT-PCR SARS-CoV-2 test during their isolation period. Of the other 100 players involved in the matches, in the following 14 days, five returned positive and 95 returned negative RT-PCR SARS-CoV-2 tests during their routine screening. All positive RT-PCR SARS-CoV-2 tests (both identified contacts and other players) were deemed to be due to internal club COVID-19 outbreaks or wider-community transmission. Despite the frequent interactions between SARS-CoV-2 positive players and other players during a rugby match, these data suggest that SARS-CoV-2 transmission is limited during this type of outdoor activity.

### Identified close contacts

The one identified contact who tested positive for SARS-CoV-2 was likely exposed to the virus within the club environment, as players from the same club who did not play in the match tested positive at the same time. In addition, transmission appears beyond the training activity, since staff members also tested positive for SARS-CoV-2. Whilst C1 was involved in 11 tackles and accumulated a high duration of close proximity with CoV1 (Figure 2B) this was similar to another player (Figure 2B; P8), not identified as a contact who did not test positive for SARS-CoV-2 in the 14-day period following the match.

CoV7 was involved in 10-14 tackles with C18, C19, C21, P7, P8, P11 (Figure 4A). None of these players tested positive for SARS-CoV-2 in the 14-days following the match, therefore if the tackle was the mechanism of transmission for C1, theoretically players from match 3 would also have tested positive for SARS-CoV-2. Both contacts (C2-C28) and other players not identified as contacts experienced a high number of tackles with SARS-CoV-2 positive players and did not test positive for SARS-CoV-2. Therefore, there appears to be limited transmission of SARS-CoV-2 during a tackle, even when directly face-to-face, as per the contact tracing framework.[16] The limited transmission may have been due to the good ventilation of an outdoor environment or minimal ‘prolonged’ face-to-face interactions during the match. P17 tested positive following match 3, was not identified as a contact and their interaction with CoV8 was only one tackle and being within 2 m for 3.8 secs (Figure 4B-C). This is an unlikely cause of transmission, given the greater interactions between SARS-CoV-2 positive and other players, who did not subsequently test positive within 14 days.

### Other observations

All match officials returned negative SARS-CoV-2 RT-PCR screening tests, after officiating matches with positive SARS-CoV-2 players. Furthermore, it appears that the ball is low risk for virus transmission given that players frequently touch the ball.

It appears that the strategy used to identify contacts, resulted in players requiring to isolate, who were not a risk, which has implications for community team sport ‘Test and Trace’ policies, impacting on a player’s ability to go to work or school. As such, participants who exceed the interactions observed in this study may be at risk of transmission, but based on the observations in this study, the transmission risk of outdoor team sports does appear low.

Outside of Super League match play, SARS-CoV-2 transmission has been observed within teams. It may be that SARS-CoV-2 transmission is greater in wider-community settings, or during indoor activities associated with team sports (e.g., gym, clubhouse, changing rooms), whereby fomite transmission is greater, and airflow is lower than outdoors, as opposed to the outdoor team sport activity *per se*. Furthermore, the risks of virus transmission events during off-field behaviours (conversations and socialising), should remain a priority for community team sports who are returning.[24]

### Study limitations

These data are limited by the small sample, and contact tracing and player interactions are limited to only match play (kick off to full time); however, players were shedding infectious virus during the match (confirmed by Ct values) and players had a high number of human-to-human interactions (confirmed by video analysis and GPS data).

Reported player interaction data provides only a snapshot of the number that likely took place between SARS-CoV-2 positive players and others. Players on the same team as SARS-CoV-2 positive players may have arrived and departed on a team coach, given all fixtures were played at away (neutral) venues, were in changing rooms pre-, half-time and post-match, and took part in a warm-up. Social distancing during non-rugby activity was advised by the governing body but it is beyond the scope of this study to determine the adherence of these protocols. Nevertheless, these findings indicate limited transmission during matches in team sports.

## Conclusion

Based on eight SARS-CoV-2 positive players participating in four rugby league matches, potentially exposing 128 other players to the virus, limited transmission was observed during the match. Positive SARS-CoV-2 observations were linked to internal club COVID-19 outbreaks or wider-community close contact transmission. Furthermore, there was no observed transmission to match officials involved in the matches. Given the return of community team sports during the COVID-19 pandemic, and potential human-to-human transmission risk during these activities, when balanced against the wider benefits of health from sport, determining the transmission risk is a priority. These are important to prevent unnecessary wider societal disruption and allow community outdoor team sports to return safely. For community team sports who may not have access to video analysis and GPS player monitoring, these data provide reassurance that even without these tools, the transmission risk during a match is likely to be very low. Further analysis of other close-contact sport settings and also the exploration of transmission risk in the training environment should be undertaken to further understand the novel findings presented.

## Data Availability

All data are presented in the manuscript.

